# Associations between general sleep quality and measures of functioning and cognition in subjects recently diagnosed with bipolar disorder

**DOI:** 10.1101/2022.07.24.22277972

**Authors:** Bruno Braga Montezano, Taiane de A. Cardoso, Luciano D. de Mattos Souza, Fernanda Pedrotti Moreira, Thaíse Campos Mondin, Ricardo A. da Silva, Karen Jansen

## Abstract

The present study aims to assess the association between sleep quality and the functional and cognitive impairment in subjects that recently converted to bipolar disorder (BD), subjects with past major depressive disorder (MDD) and subjects with recurrent MDD. This was a cross-sectional study corresponding to a second wave of a cohort study with a community sample. The first wave included 585 subjects diagnosed with MDD. Mood episodes were assessed through structured clinical interview. Functional and objective cognitive impairments were measured by the Functional Assessment Short Test (FAST) and Letter-number sequencing from Wechsler Adult Intelligence Scale III, respectively, and Cognitive Complaints in Bipolar Disorder Rating Assessment (COBRA) was used for subjective cognition measure. Sleep quality was assessed by Pittsburgh Sleep Quality Index (PSQI). In PSQI, FAST and COBRA, we found significantly worse scores in the BD and recurrent MDD groups when compared to past MDD group. Our findings also showed a significant association between functioning and subjective cognition with general sleep quality in all observed groups. We reinforce the need to follow-up for maintenance of functional and cognitive impairment, notably with BD patients, who may suffer in addition to damage caused by sleep alterations, also with neuroprogression in the long term.

## 1. Introduction

Bipolar disorder (BD) is a severe and chronic psychiatric disorder, characterized by depressive, (hypo)manic and/or mixed episodes (McIntyre et al., 2020). Sleep alterations are characteristics in both depressive and manic or hypomanic episodes (Ritter et al., 2015; Slyepchenko et al., 2019). Impairments related to sleep patterns are also perceived in patients in a period of euthymia or remission from the episode (De la Fuente-Tomás et al., 2018).

Some studies suggest that mood disorders are associated with poorer functioning and cognitive performance when compared to healthy controls, especially BD (Reyes et al., 2017; Kapczinski et al., 2016). Studies that have been examining the effect of sleep disturbances on functioning have found greater functional impairment in subjects with sleep problems (Lai et al., 2014). In addition, worse sleep can predict greater impairment in functioning (Slyepchenko et al., 2019). As in functioning, studies show that a worse sleep is associated to a worst cognitive performance (Russo et al., 2015; Kaplan, 2020). Moreover, sleep-related alterations, such as sleep variability, may predict poor working memory and verbal learning performance (Kanady et al., 2017).

Most of the studies evaluate late samples of subjects diagnosed with BD. Therefore, the effects of impaired sleep on functioning and cognition may be mistakenly assessed due to the impact of disease neuroprogression (Kapczinski et al., 2017). Furthermore, most studies in the literature that assessed cognition in samples of subjects with mood disorders considered objective measures. This study seeks to evaluate the construct both objectively and subjectively, considering the possible inconsistencies between the criteria.

Given this, it is important that further studies can verify if there are effects of sleep on functional and cognitive impairment in samples recently diagnosed with BD, or if these implications are shown only in later and impaired samples due to the progression of the disorder. Considering these aspects, the present study aims to assess the association between sleep quality and the functional and cognitive impairment in subjects that recently converted to BD, subjects with past major depressive disorder (MDD) and subjects with recurrent MDD.

## 2. Methods

This was a cross-sectional study corresponding to a second wave of a cohort study with a community sample. The first wave included 966 participants, between the ages of 18 to 60 years, living in the urban area of Pelotas, RS (Brazil) in the period from 2012 to 2015, in which 585 of them were diagnosed with MDD.

Data of the second wave were collected from 2017 to 2018; specifically, at a mean of three years after baseline, all subjects that were diagnosed with MDD were invited to participate in a reassessment (n = 468). All subjects were informed about the research objectives, and they signed an informed consent document. The respondents who were diagnosed with psychiatric disorder in the clinical interview during the first or second wave were referred to treatment at the Clinic of Research and Extension in Mental Health of the Catholic University of Pelotas (UCPel). This study was approved by the Research Ethics Committee of UCPel under protocol number 502.604.

The Mini International Neuropsychiatric Interview (MINI) has been administered by trained psychologists to diagnose mood disorders. The MINI is a short questionnaire organized by independent disorders modules, following the criteria of DSM-IV and ICD-10 that can be administrated after a brief training (Sheehan et al., 1998). In case of diagnosis doubt, the patients were reassessed by a experienced psychiatrist to confirm or refute the diagnosis. We only used data from the clinical interview of the second wave. Thus, the subjects were divided into three groups: (1) subjects that converted their diagnosis to BD; (2) subjects with recurrent major depressive episode; (3) subjects that did not fill criteria for current major depressive or (hypo)manic episode. However, the subjects in the third group could have another psychiatric diagnose.

For the assessment of general sleep quality, we used the Pittsburgh Sleep Quality Index (PSQI), that is an instrument structured by nineteen self-assessed by the subject and five questions answered by the room or bed partner. The total score can range from 0 to 21, considering that a higher score means a worse general sleep quality (Bertolazi et al., 2011). To assess the global functioning, the Functional Assessment Short Test (FAST) was used, which consists of 24 items built to assess six areas of functioning: autonomy, work, cognition, finances, interpersonal relationships and leisure (Cacilhas et al., 2009). The sum of all items results in an overall score, called global functioning, where the higher the score, highter the functional impairment. The Cognitive Complaints in Bipolar Disorder Rating Assessment (COBRA) was used to assess subjective perception of cognition validated to Brazil by Lima et al. (2018). COBRA consists of sixteen self-reported items, formed by the following domains: executive functioning, processing speed, working memory, memory and verbal learning, attention/concentration and mental tracking. It is scored within a range of 0-48, with higher scores indicating a greater number of cognitive disorders. The cognitive performance was assessed using the Sequence of Numbers and Letters from Wechsler Adult Intelligence Scale III (WAIS-III). The instrument measures working memory, mental manipulation, attention, concentration and short-term auditory memory. The subtest consists of seven items, and each item has three trials, resulting in a total score when added up (Wechsler, 2004).

The data were collected through the Open Data Kit Collect Software in version 1.1.7, on tablets, and later transferred to spreadsheets. For data analysis, scripts were written in the R programming language, version 4.0.3 (R Core Team, 2021). Categorical variables will be described in absolute and relative frequencies. Numerical variables will have their distribution tested on the Gauss curve and will be presented by mean and standard deviation or median and interquartile intervals, according to its distribution. The correlation between sleep quality and measures of functioning and cognition will be tested by Pearson or Spearman, if necessary a model of analysis by linear regression will be constructed to adjust for confounding variables. Variables associated with exposure and outcome with p*<*0.20 in the crude analysis will be considered possible confounding factors. In order to check median differences among groups, the Kruskal-Wallis test will be used. Subsequently, in case of a significant Kruskal-Wallis test, Dunn test and Dwass-Steele-Critchlow-Fligner test were used as post-hoc. The hyphotesis tests were considered statistically significant when p*<*0.05.

## 3. Results

Out of the eligible sample (n = 585), 20% could not be located or refused to participate in the second wave. Thus, the current sample included 468 patients. Fifty-eight subjects were included in the BD group, one hundred forty-nine subjects in the recurrent MDD group, and two hundred and sixty-one subjects in the past MDD group. Sociodemographic and clinical characteristics are shown in Table 1 and Table 2.

**Table 1:** Sociodemographic characteristics across the groups.

| Variables | BD<br>(n = 58) | Past MDD<br>(n = 261) | Recurrent<br>MDD<br>(n = 149) | p-value |
| --- | --- | --- | --- | --- |
| **Sex** |  |  |  | 0.040 |
| Female | 55 (94.8%) | 213 (81.6%) | 127 (85.2%) |  |
| Male | 3 (5.2%) | 48 (18.4%) | 22 (14.8%) |  |
| **Age** | 37.40 ($\pm 10.73$) | 40.69 ($\pm 11.48$) | 41.00 ($\pm 11.22$) | 0.142 |
| **Ethnicity** |  |  |  | 0.019 |
| White | 37 (63.8%) | 208 (80.0%) | 107 (72.3%) |  |
| Not white | 21 (36.2%) | 52 (20.0%) | 41 (27.7%) |  |
| **Socioeconomic status** |  |  |  | 0.082 |
| A/B (upper) | 18 (31.0%) | 117 (45.0%) | 55 (37.2%) |  |
| C/D/E (lower) | 40 (69.0%) | 143 (55.0%) | 93 (62.8%) |  |
| **Years of education** | 9.40 ($\pm 4.25$) | 10.95 ($\pm 3.65$) | 9.86 ($\pm 3.32$) | <0.001 |
| **Current occupation** |  |  |  | 0.038 |
| Working and/or study | 38 (65.5%) | 173 (66.5%) | 80 (54.1%) |  |
| None | 20 (34.5%) | 87 (33.5%) | 68 (45.9%) |  |
| **Marital status** |  |  |  | 0.272 |
| Has a partner | 31 (53.5%) | 169 (64.8%) | 94 (63.1%) |  |
| Does not have a partner | 27 (46.6%) | 92 (35.3%) | 55 (36.9%) |  |

**Table 2:** Clinical characteristics across the groups.

| Variables | BD<br>(n = 58) | Past MDD<br>(n = 261) | Recurrent<br>MDD<br>(n = 149) | p-value |
| --- | --- | --- | --- | --- |
| **Alcohol**<br>**abuse/dependence** |  |  |  | 0.254 |
| No | 38 (67.9%) | 178 (73.5%) | 111 (78.7%) |  |
| Yes | 18 (32.1%) | 64 (26.5%) | 30 (21.3%) |  |
| **Illicit substance**<br>**abuse/dependence** |  |  |  | 0.418 |
| No | 37 (66.1%) | 175 (72.3%) | 94 (66.7%) |  |
| Yes | 19 (33.9%) | 67 (27.7%) | 47 (33.3%) |  |
| **Hypnotics/sedatives**<br>**abuse/dependence** |  |  |  | 0.307 |
| No | 40 (71.4%) | 185 (76.5%) | 98 (69.5%) |  |
| Yes | 16 (28.6%) | 57 (23.5%) | 43 (30.5%) |  |
| **Current suicide risk** |  |  |  | <0.001 |
| No | 25 (43.1%) | 153 (58.6%) | 60 (40.3%) |  |
| Yes | 33 (56.9%) | 108 (41.4%) | 89 (59.7%) |  |
| **Lifetime psychiatric**<br>**medication** |  |  |  | 0.004 |
| No | 6 (10.5%) | 56 (22.5%) | 16 (10.8%) |  |
| Yes | 51 (89.5%) | 193 (77.5%) | 132 (89.2%) |  |
| **PSQI** | 10 (6.0, 14.0) | 6 (4.0, 9.0) | 12 (9.0, 14.0) | <0.001 |
| **FAST** | 25 (13.3, 35.0) | 11 (5.0, 19.0) | 29 (19.0, 39.0) | <0.001 |
| **COBRA** | 19.5 (10.3, 26.0) | 8 (5.0, 14.0) | 21 (13.0, 29.5) | <0.001 |
| **Letter-number**<br>**sequencing test** | 6 (4.0, 9.0) | 7 (5.0, 9.0) | 5 (4.0, 9.0) | 0.073 |

In PSQI, FAST and COBRA, significant score differences were found between the groups. After post-hoc, the tests found differences between the past MDD group when compared to the BD (p*<*0.001) and recurrent MDD (p*<*0.001) groups. No difference was found in Letter-number sequencing score (p=0.073).

In BD group, there was a positive correlation between PSQI and COBRA scores (r=0.636, p*<*0.001) and between PSQI and FAST scores (r=0.687, p*<*0.001), and there was a negative correlation between PSQI and WAIS subtest scores (r=-0.366, p=0.005). In recurrent MDD group, there was a positive correlation between PSQI and COBRA scores (r=0.236, p=0.004) and between PSQI and FAST scores (r=0.394, p*<*0.001), and there was no correlation between PSQI and WAIS subtest (r=-0.016, p=0.852). In past MDD group, there was a positive correlation between PSQI and COBRA scores (r=0.497, p*<*0.001) and between PSQI and FAST scores (r=0.468, p*<*0.001), and no correlation was found between PSQI and WAIS subtest scores (r=-0.004, p=0.955) in that group.

Table 3 shows the crude and adjusted analysis of general sleep quality associated with global functioning, subjective cognition and cognitive performance between the diagnosis groups. The FAST scores (global functioning) were associated with general sleep quality in all three groups. After adjusting for years of education, lifetime psychiatric medication use, and hypnotics abuse/dependence in BD group, the association was mantained (p*<*0.001). The association was also mantained in the recurrent MDD group after the adjusted analysis (p*<*0.001), considering years of education and current suicide risk as possible confounding factors.

**Table 3:** Crude and adjusted analysis of general sleep quality score (PSQI) associated with functioning and cognition between diagnose groups.

| **FAST** |  |  |  |  |
| --- | --- | --- | --- | --- |
| **Crude analysis** |  | **Adjusted analysis** |  |  |
| **B (95% CI)** | **p-value** | **B (95% CI)** | **p-value** |  |
| **BD** | 2.17 (1.60 – 2.75) | <0.001 | 1.78 (1.17 – 2.38)<sup>a</sup> | <0.001 |
| **Recurrent MDD** | 1.49 (0.96 – 2.03) | <0.001 | 1.29 (0.73 – 1.85)<sup>b</sup> | <0.001 |
| **Past MDD** | 1.48 (1.33 – 1.82) | <0.001 | — | — |
| **COBRA** |  |  |  |  |
| **BD** | 1.40 (0.97 – 1.84) | <0.001 | 1.22 (0.78 – 1.65)<sup>c</sup> | <0.001 |
| **Recurrent MDD** | 0.71 (0.28 – 1.13) | 0.001 | 0.47 (0.04 – 0.91)<sup>b</sup> | 0.033 |
| **Past MDD** | 1.08 (0.82 – 1.33) | <0.001 | — | — |
| **Letter-number sequencing** |  |  |  |  |
| **BD** | −0.26 (−0.44 – −0.09) | 0.003 | −0.11 (−0.26 – 0.05)<sup>d</sup> | 0.177 |
| **Recurrent MDD** | −0.03 (−0.17 – 0.11) | 0.667 | 0.06 (−0.06 – 0.19)<sup>c</sup> | 0.319 |
| **Past MDD** | −0.02 (−0.14 – 0.09) | 0.659 | — | — |

<sup>a</sup> Adjusted for years of education, lifetime psychiatric medication use, and hypnotics abuse/dependence.
<sup>b</sup> Adjusted for years of education and current suicide risk.
<sup>c</sup> Adjusted for years of education.
<sup>d</sup> Adjusted for years of education and hypnotics abuse/dependence.
**Legend:** Bipolar Disorder (BD); Major Depressive Disorder (MDD); Functioning Assessment Short Test (FAST); Cognitive Complaints in Bipolar Disorder Rating Assessment (COBRA).

COBRA scores (subjective cognition) were also associated with general sleep quality in all groups. In BD group, after adjusting for years of education, the association remained (p*<*0.001). In recurrent MDD group, association was mantained (p=0.033), after adjusting for years of education and current suicide risk.

WAIS subtest scores (cognitive performance) were associated with general sleep quality only in BD group (p=0.003). However, after adjusting for years of education and hypnotics abuse/dependence, the association between cognitive performance and sleep quality in subjects with BD was not mantained (Table 3).

## 4. Discussion

The present study investigated the association between sleep quality and measures of functioning and cognition in subjects recently diagnosed with BD, and compared those measures with two MDD groups. Our main findings include greater functional impairment and cognitive complaints (subjective cognition) in BD and recurrent MDD groups when compared with past MDD group. Our findings also showed an significant association between global functioning and subjective cognition with general sleep quality in all observed groups.

There is evidence in the literature of worse sleep quality in subjects with mood disorders when compared to healthy subjects (Lai et al., 2014; Boland et al., 2015; Bradley et al., 2017; Slyepchenko et al., 2019). Furthermore, there are studies, such as Schneider et al. (2008); Bo et al. (2019), that indicate worse cognitive performance in BD and MDD patients when compared to healthy controls (HC). It is worth mentioning that our findings are supported by some other studies that demonstrated sleep disturbances primarily associated with cognitive performance in BD patients (Rosa et al., 2013; Russo et al., 2015). Our results are consistent with a study by Volkert et al. (2015), which showed that the more sleep disturbances the patients reported, the worse they performed on cognitive tests.

Given this, we found that cognitive impairment was greater in subjects with recurrent MDD and in the group that converted to BD. In our sample, patients with worse sleep quality demonstrated worse subjective cognition and cognitive performance, despite cognitive performance have not mantained the association. These findings can also be interpreted from the perspective of the potential impact of cognitive impairment and poor sleep quality observed in some patients – characteristics considered necessary to achieve proper functioning and to engage in social and work activities in general. However, Torres et al. (2020) observed that properly treated patients with BD showed a cognitive improvement in the first three years after an initial manic episode giving evidence against the effect of neuroprogression at this stage of the disease.

Next we explored the relationship between sleep quality and functioning and found, as might be expected, that worse sleep quality were associated with poorer functioning. Regarding this, the results of this study corroborate previous findings that showed functional impairment being predicted by sleep measures (Slyepchenko et al., 2019). Studies that measure functioning indicate greater functional impairment in subjects with mood disorders, especially BD, when compared to HC (Kapczinski et al., 2016; Reyes et al., 2017). Prior work suggests that sleep disturbances predicted greater functional impairment, similar to the results of this study (Walz et al., 2013). In the present study, comparisons were not made with HC, but instead with subjects who had experienced past MDD.

The design of the cross-sectional study prevents the inference of the causal relationship between sleep quality and measures of cognition and functioning. Cohort studies are needed to provide stronger evidence and establish whether such relationships exist. Strengths of the present study include the fact that patients with BD have been recently diagnosed, developing the disorder in the last three years, reducing the possible effect of neuroprogression on the clinical condition.

In conclusion, we demonstrated an association between general sleep quality and global functioning and cognition, among patients with BD and recurrent MDD especially. Moreover, the study suggests the need to follow-up for the maintenance of functional and cognitive impairment, notably with BD patients, who may suffer in addition to damage caused by sleep alterations, also with the disease neuroprogression in the long term. Neuroprogression is associated with structural and functional changes in the brain that could trigger even more functional and cognitive impairment. On top of that, illness neuroprogression may therefore lead to hospitalizations, worse quality of life, shortening of inter-episodic intervals and possibily suicide attempts. This shows even greater importance for attention to residual symptoms and brain health maintance in treatments aimed at improving the clinical condition of BD patients.

## Data Availability

Research data are not shared due to privacy and anonymity reasons outlined by the authors of the original research project. The analysis scripts can be accessed through the link: https://github.com/brunomontezano/sleep-quality-cognition

